# Real-world Effectiveness and Causal Mediation Study of BNT162b2 on Long COVID Risks in Children and Adolescents

**DOI:** 10.1101/2024.02.19.24302823

**Authors:** Qiong Wu, Bingyu Zhang, Jiayi Tong, L. Charles Bailey, H. Timothy Bunnell, Jiajie Chen, Elizabeth A. Chrischilles, Dimitri A. Christakis, Stephen M. Downs, Kathryn Hirabayashi, Ravi Jhaveri, Aaron D. Mishkin, Abu S.M. Mosa, Nathan M. Pajor, Suchitra Rao, Hanieh Razzaghi, Hayden T. Schwenk, Marion R. Sills, Huiyuan Wang, Linbo Wang, Yudong Wang, Dazheng Zhang, Ting Zhou, Eric J. Tchetgen Tchetgen, Jeffrey S. Morris, Christopher B. Forrest, Yong Chen the RECOVER Consortium

## Abstract

**Background:** The impact of pre-infection vaccination on the risk of long COVID remains unclear in the pediatric population. Further, it is unknown if such pre-infection vaccination can mitigate the risk of long COVID beyond its established protective benefits against SARS-CoV-2 infection.

**Objective:** To assess the effectiveness of BNT162b2 on long COVID risks with various strains of the SARS-CoV-2 virus in children and adolescents, using comparative effectiveness methods. To disentangle the overall effectiveness of the vaccine on long COVID outcomes into its independent impact and indirect impact via prevention of SARS-CoV-2 infections, using causal mediation analysis.

**Design:** Real-world vaccine effectiveness study and mediation analysis in three independent cohorts: adolescents (12 to 20 years) during the Delta phase, children (5 to 11 years) and adolescents (12 to 20 years) during the Omicron phase.

**Setting:** Twenty health systems in the RECOVER PCORnet electronic health record (EHR) Program.

**Participants:** 112,590 adolescents (88,811 vaccinated) in the Delta period, 188,894 children (101,277 vaccinated), and 84,735 adolescents (37,724 vaccinated) in the Omicron period.

**Exposures:** First dose of the BNT162b2 vaccine vs. no receipt of COVID-19 vaccine.

**Measurements:** Outcomes of interest include conclusive or probable diagnosis of long COVID following a documented SARS-CoV-2 infection, and body-system-specific condition clusters of post-acute sequelae of SARS-CoV-2 infection (PASC), such as cardiac, gastrointestinal, musculoskeletal, respiratory, and syndromic categories. The effectiveness was reported as (1-relative risk)*100 and mediating effects were reported as relative risks.

**Results:** During the Delta period, the estimated effectiveness of the BNT162b2 vaccine against long COVID among adolescents was 95.4% (95% CI: 90.9% to 97.7%). During the Omicron phase, the estimated effectiveness against long COVID among children was 60.2% (95% CI: 40.3% to 73.5%) and 75.1% (95% CI: 50.4% to 87.5%) among adolescents. The direct effect of vaccination, defined as the effect beyond their impact on SARS-CoV-2 infections, was found to be statistically non-significant in all three study cohorts, with estimates of 1.08 (95% CI: 0.75 to 1.55) in the Delta study among adolescents, 1.24 (95% CI: 0.92 to 1.66) among children and 0.91 (95% CI: 0.69 to 1.19) among adolescents in the Omicron studies. Meanwhile, the estimated indirect effects, which are effects through protecting SARS-CoV-2 infections, were estimated as 0.04 (95% CI: 0.03 to 0.05) among adolescents during Delta phase, 0.31 (95% CI: 0.23 to 0.42) among children and 0.21 (95% CI: 0.16 to 0.27) among adolescents during the Omicron period.

**Limitations:** Observational study design and potentially undocumented infection.

**Conclusions:** Our study suggests that BNT162b2 was effective in reducing risk of long COVID outcomes in children and adolescents during the Delta and Omicron periods. The mediation analysis indicates the vaccine’s effectiveness is primarily derived from its role in reducing the risk of SARS-CoV-2 infection.

**Primary Funding Source:** National Institutes of Health.

## Introduction

The scientific and clinical understanding of the long-term effects or sequelae following a COVID-19 infection caused by the SARS-CoV-2 virus continues to evolve. These post-acute sequelae of SARS-CoV-2 infection (PASC), also referred to as long COVID, are persistent, exacerbated, or newly developed symptoms or other health effects that can affect multiple organ systems (e.g., cardiovascular, neurologic, mental, metabolic, and renal)^1–6^. Significant work has been focused on characterizing the complex clinical representation of PASC with the development of standardized definitions. Research concerning PASC in pediatric populations revealed a difference in clinical features and incidence rates compared to adults^7^.

The effectiveness of COVID-19 vaccines in preventing symptomatic and severe COVID-19 has been assessed through randomized controlled trials (RCTs^8,9^) and subsequent observational vaccine-effectiveness studies^10–17^. However, our understanding of how a COVID-19 vaccine administered prior to infection impacts long COVID outcomes is still unclear. Further, research conducted to date has been mainly centered on adults and has produced inconsistent findings. Some studies suggest a significant protective effect^18–27^, e.g., a reduced risk of the diagnosis of PASC or experiencing certain PASC symptoms. Meanwhile, other studies indicate mixed effects revealing considerable variations across different age groups, various dominant virus strains, and distinct PASC symptoms^28–32^, or even suggesting counter-protective effects^33,34^. Moreover, the majority of existing studies have reported effects by comparing breakthrough infection to infections in unvaccinated individuals, which is conditional on the infection status^18,21,24,26,28,29,35^. This approach only reveals the vaccine’s effectiveness within the infected population, which does not accurately represent the true impact of vaccination on long COVID, as the risk of infection is substantially reduced in the vaccinated group^10–17^. Furthermore, this approach, conditioning on post-treatment variables such as infection status, could introduce selection bias, as underscored by Hernan et al. (2023)^36^.

To address these gaps in our knowledge of the effectiveness of COVID-19 vaccination on long COVID outcomes among the understudied pediatric population, we designed this study among children and adolescents during the Delta and Omicron variant-predominant periods using electronic health record (EHR) data from RECOVER PCORnet Program ^37,38^. It is, to the best of our knowledge, among the first studies in the U.S. focusing on studying the effectiveness of COVID-19 vaccination on long COVID within the pediatric demographic. Further, to provide a comprehensive understanding of the vaccine’s effect and mitigate the potential bias from conditioning on the post-treatment infection, we conducted a causal mediation analysis^39–41^ that quantifies both the overall vaccine effectiveness and effects through specific mediating pathways. The overall vaccine effectiveness yields a quantification of the impact on long COVID involving both infected and uninfected individuals, not only infected patients, which delivers more generalizable findings. Meanwhile, the dissection of the overall vaccine effectiveness into direct and indirect components through causal mediation analysis allows for a nuanced assessment of the vaccine’s influence on long COVID outcomes, *either beyond or through* the prevention of SARS-CoV-2 infections. To strengthen the reliability of our research findings, we also conducted the proximal analysis utilizing a set of negative control exposures and outcomes^42,43^, which helped to assess the potential residual bias due to unmeasured confounders in the EHR data.

## Methods

### Data sources

This study is part of the NIH Researching COVID to Enhance Recovery (RECOVER) Initiative, which seeks to understand, treat, and prevent PASC. For more information on RECOVER, visit https://recovercovid.org/. Participating institutions in this study included: Cincinnati Children’s Hospital Medical Center, Children’s Hospital of Philadelphia, Children’s Hospital of Colorado, Duke University, University of Iowa Healthcare, Ann & Robert H. Lurie Children’s Hospital of Chicago, Medical College of Wisconsin, University of Michigan, University of Missouri, Medical University of South Carolina, Nationwide Children’s Hospital, Nemours Children’s Health System (in Delaware and Florida), OCHIN, Inc, Ohio State University, Seattle Children’s Hospital, Stanford University, Temple University, University of California San Francisco, Vanderbilt University Medical Center, and Wake Forest Baptist Health. Data were transformed to either the PCORnet or Observational Medical Outcomes Partnership (OMOP) data models^44,45^, and forwarded to the RECOVER-PCORnet Coordinating Center. For this study, we used the s9 version of the data, collected till June 2023, which comprises 6,868,813 patients.

### Construction of study cohorts

We identified three study cohorts to assess both the effectiveness and mediating effects of the BNT162b2 vaccine on long COVID risks associated with various strains of the SARS-CoV-2 virus in children and adolescents in the United States. Study 1 involved adolescents focused on the period when the Delta variant was prevalent, specifically, from July 1, 2021, to November 30, 2021. Study 2 involved children and Study 3 among adolescents covered a period when the Omicron variant was prevalent from January 1 to November 30, 2022, ensuring a 179-day follow-up period for the observation of long COVID outcomes.

To be eligible for the study, children had to be aged between 5 and 11 years, while adolescents were between 12 and 20 years old at the start of each study. Participants could not have received a COVID-19 vaccination or had a documented SARS-CoV-2 infection at the start of the study period. Moreover, to confirm their active engagement with the healthcare system, participants must have had a prior interaction (either in-person, via phone, or through telehealth) within the 18 months leading up to their cohort entry. To ensure sufficient follow-up time for documenting infections among participants, individuals in the study cohorts were required to enroll at least three months before the end of the study for the Delta variant period, and at least four months before the end of the Omicron variant period.

The selection of participants in three study cohorts is summarized in an attrition table (i.e., Figure 1). A detailed description of the cohort construction and observation windows is available in the Supplemental Appendix Section S1.

**Figure 1.**
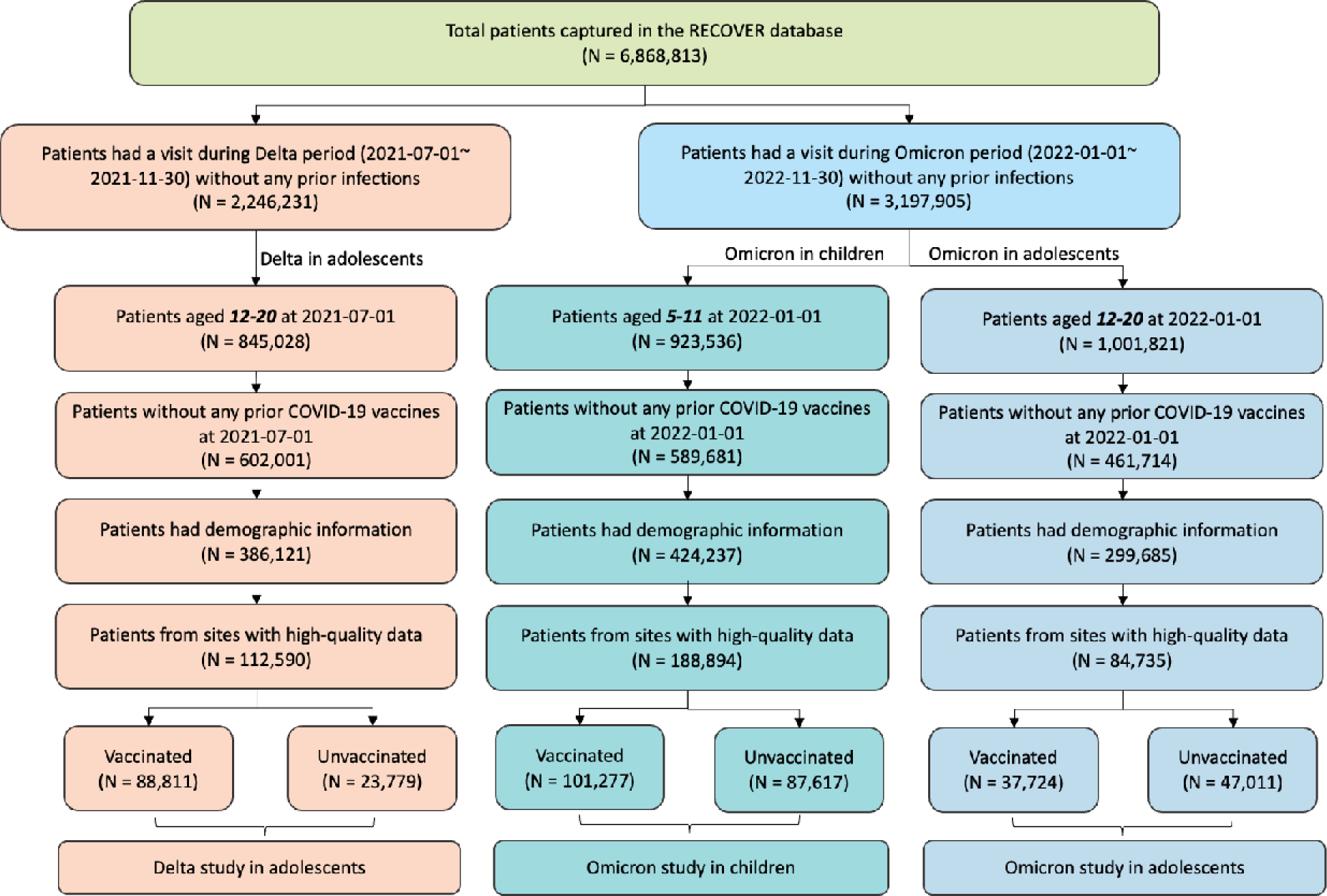
Selection of participants for the three study cohorts evaluating the overall effectiveness, direct and indirect effect of the BNT162b2 vaccine on long COVID outcomes in (1) adolescents aged 12-20 years during the period when the Delta variant was prevalent, (2) children aged 5 to 11 years and (3) adolescents aged 12 to 20 years during the period when the Omicron variant was prevalent.

### Study variables

#### Intervention

The intervention of interest was vaccination, in comparison with no receipt of any type of COVID-19 vaccine. In our database, the vaccination records for children and adolescents showed 89.3% for BNT162b2, 2.1% for mRNA-1273, and 8.6% for unspecified COVID-19 vaccines. Considering that the BNT162b2 vaccine constituted the majority of recorded vaccinations for this age group in our database, our study mainly focused on evaluating the BNT162b2 vaccine. Nevertheless, a comprehensive sensitivity analysis covering all types of COVID-19 vaccines available in the U.S. can be found in the Supplementary Appendix Section S8.

We defined the index date for the intervention group as the date of receiving the first dose of the BNT162b2 vaccine. For the comparator group, an index date was assigned based on a randomly chosen medical visit while ensuring the distribution of index dates for the comparator group matched the distribution in the vaccination group to control for temporal effects.

#### Mediator

The mediator in our study was identified as any documented SARS-CoV-2 infections defined by the occurrence of positive polymerase-chain-reaction (PCR), serology, or antigen tests or diagnoses of COVID-19, post-acute sequelae of SARS-CoV-2 (PASC), or multisystem inflammatory syndrome (MIS) regardless of the presence of symptoms. We defined the risk period of infections as 28 days after the index date such that participants with infections within 28 days were excluded.

#### Outcomes

The primary outcome in our study was a conclusive or probable diagnosis of long COVID 28 to 179 days following a documented SARS-CoV-2 infection. A conclusive diagnosis of long COVID was determined by two or more medical visits indicating diagnoses of PASC or MIS. Recognizing the potential limitations and varying uptake of specific long-COVID diagnostic codes, we also identified a probable diagnosis of long COVID. This was characterized by a single visit indicating a diagnosis of PASC or MIS or the presence of a documented SARS-CoV-2 infection alongside a minimum of two long-COVID-compatible diagnoses, spaced at least 28 days apart. These long-COVID-compatible diagnoses were categorized based on previously identified clusters of codes associated with post-acute manifestations of COVID-19 in earlier research^7,46^. In addition, we identified body-system-specific PASC condition clusters, including cardiac, gastrointestinal, musculoskeletal, respiratory, and syndromic categories, as secondary outcomes given the heterogeneity of long COVID symptoms. The diagnostic criteria for these body-system-specific PASC clusters required a documented SARS-CoV-2 infection with at least two relevant diagnoses from the respective cluster, spaced a minimum of 28 days apart.

#### Confounding variables

To account for potential confounding in the relation of intervention, mediator, and outcome, an extensive set of confounding variables was incorporated including demographic variables such as age, sex, and race/ethnicity; clinical factors like obesity status, a chronic condition indicator as defined by the Pediatric Medical Complexity Algorithm (PMCA), and a list of pre-existing chronic conditions; and healthcare utilization metrics including the number of inpatient and outpatient visits, emergency department (ED) visits, unique medications prescribed, and the count of negative COVID-19 tests administered prior to the cohort entry. The detailed definitions of study variables were included in the Table S1 of the Supplementary Appendix.

### Statistical analysis

We estimated the overall vaccine effectiveness and mediating effects of BNT162b2 vaccines on long COVID risks by conducting causal mediation analyses with documented infection as a mediating variable. A visual representation of the hypothesized effect pathway can be found in Figure S1 of the Supplementary Appendix. The overall vaccine effectiveness was quantified by the total effect of the mediation analysis which represents a marginal effect of vaccination on the risk of long COVID. We further decomposed the overall vaccine effectiveness (total effect) into (natural) direct and indirect effects, to determine the roles of BNT162b2 vaccines in preventing long COVID outcomes. Direct effects measure the impact of vaccination on long COVID outcomes *beyond* its effect on COVID-19 infection, while indirect effects capture its impact *through* prevention of infection.

The overall vaccine effectiveness (total effect), and direct and indirect effects were estimated by implementing the weighting strategy to adjust for a large number of confounding variables. For each study cohort, we used propensity scores to mimic the treatment assignment in randomized experiments^47,48^ and mediator probabilities to adjust for confounding in the mediator-outcome relationship^49,50^. For each study cohort, three logistic regression models were built to estimate (1) the propensity score as the probability of a participant belonging to the vaccination group, (2) mediator probability in the vaccination group as the probability of a participant in the vaccination group being infected during the follow-up period, and (3) mediator probability in the comparator group as the probability of a participant in the comparator group being infected during the follow-up period. We trimmed participants based on 5^th^ and 95^th^ percentile cutpoints of the propensity scores and mediator probabilities to stabilize weights and improve robustness^51^. We derived and implemented closed-form variance estimators for overall vaccine effectiveness (total effect), and direct and indirect effects using the empirical sandwich method^52^. Covariate balance is assessed using the standardized mean difference (SMD) before and after weighting, with an SMD value below 0.1 indicating an acceptable balance^53^.

The overall vaccine effectiveness was reported as (1-relative risk)*100. The direct and indirect effects of the three study cohorts are reported in relative risks (change in likelihood). The corresponding risk differences (change in incidence) are presented in Figure S3 of the Supplementary Appendix for the straightforward interpretations.

### Sensitivity analysis

We conducted a comprehensive set of sensitivity analyses to evaluate the robustness of the research findings. Sections S5-9 of the Supplementary Appendix present sensitivity analyses for the impacts of cohort design. We present the overall vaccine effectiveness and mediation effects for both Delta and Omicron studies focused on adolescents aged 12 to 17 years. In the Delta study among adolescents, the diagnosis of long COVID was defined based on 28 to 179 days following an infection in the Delta period, which could exceed the Delta period. We acknowledge the possibility of subsequent Omicron infections following an initial Delta infection prior to the diagnosis of long COVID. To ensure that our study specifically addresses long COVID as a consequence of Delta variant infections, we have narrowed the observation window for long COVID to the Delta-dominant period in a sensitivity analysis. Since the vaccination group had a notably lower percentage of patients entering through ED visits compared to the comparator group, we carried out a sensitivity analysis excluding those who joined the cohort due to an ED visit. To examine the dose-dependent effects of the BNT162b2 vaccine, we estimated the effects of a two-dose vaccination which was defined as the administration of a second dose at least 14 days before the infection. Additionally, we provided estimates of the vaccine’s effects including all available COVID-19 vaccine brands.

Section S4, and S10-11 of the Supplementary Appendix focused on sensitivity analyses regarding the statistical methodologies employed in the study. Beyond our primary causal mediation weighting, we present findings on the overall vaccine effectiveness and direct and indirect effects using a regression-based approach for causal mediation analysis. It’s noted that the measured confounding variables may not be sufficiently rich to account for confounding. Hence, we conducted the proximal analysis using 4 pre-specified negative control variables from pediatric physicians which are believed to be not causally related to the intervention, mediator, and outcome^42,43^.

## Results

### Study population

Within the RECOVER network, we identified a total of 112,590 adolescents in which to investigate both the overall vaccine effectiveness and mediating effects of BNT162b2 on long COVID outcomes during a period dominated by the Delta variant (refer to Table 1 for baseline characteristics). For studying the overall vaccine effectiveness and mediating effects of BNT162b2 on the risk of long COVID of Omicron infections, the cohort comprised 188,894 children and 84,735 adolescents (baseline characteristics provided in Table 2). The highest incidence rate of both the long COVID outcome and documented infection was observed in the unvaccinated group of the Delta study in adolescents (i.e., 3.54 and 53.10 per 10,000 person-week). In contrast, the vaccinated group from the same study exhibited the lowest incidence rates, being 0.11 and 1.97 per 10,000 person-weeks, respectively. Across all three cohorts, there was a minor imbalance in testing rates prior to cohort entry between the vaccinated and unvaccinated groups. However, after applying propensity score adjustments, all covariates achieved the balance between the vaccinated and unvaccinated groups, evidenced by an SMD of less than 0.1 in all three cohorts (Figures S2-4 of the Supplementary Appendix).

**Table 1.** Baseline characteristics of adolescents 12 to 20 years of age in the study of overall effectiveness, direct and indirect effect of the BNT162b2 vaccine on long COVID during the period when the Delta variant was prevalent.

| *Delta study in adolescents* |  |  |  |
| --- | --- | --- | --- |
|  | **Vaccinated**<br>**(N=88,811)** | **Unvaccinated**<br>**(N=23,779)** | **Overall**<br>**(N=112,590)** |
| **Age** |  |  |  |
| **Median [Q1, Q3]** | 14 [13, 16] | 15 [13, 17] | 15 [13, 16] |
| **Distribution** |  |  |  |
| **12** | 16,867 (19.0%) | 3,331 (14.0%) | 20,198 (17.9%) |
| **13** | 15,697 (17.7%) | 3,107 (13.1%) | 18,804 (16.7%) |
| **14** | 15,127 (17.0%) | 3,214 (13.5%) | 18,341 (16.3%) |
| **15** | 14,204 (16.0%) | 3,138 (13.2%) | 17,342 (15.4%) |
| **16** | 10,091 (11.4%) | 2,985 (12.6%) | 13,076 (11.6%) |
| **17** | 7,580 (8.5%) | 2,771 (11.7%) | 10,351 (9.2%) |
| **18** | 4,098 (4.6%) | 2,043 (8.6%) | 6,141 (5.5%) |
| **19** | 2,936 (3.3%) | 1,778 (7.5%) | 4,714 (4.2%) |
| **20** | 2,211 (2.5%) | 1,412 (5.9%) | 3,623 (3.2%) |
| **Gender** |  |  |  |
| **Female** | 46,650 (52.5%) | 12,830 (54.0%) | 59,480 (52.8%) |
| **Male** | 42,161 (47.5%) | 10,949 (46.0%) | 53,110 (47.2%) |
| **Ethnicity** |  |  |  |
| **White** | 26,971 (30.4%) | 12,373 (52.0%) | 39,344 (34.9%) |
| **Black/AA** | 16,044 (18.1%) | 4,833 (20.3%) | 20,877 (18.5%) |
| **Hispanic** | 35,147 (39.6%) | 4,112 (17.3%) | 39,259 (34.9%) |
| **Asian/PI** | 3,521 (4.0%) | 426 (1.8%) | 3,947 (3.5%) |
| **Multiple** | 1,521 (1.7%) | 520 (2.2%) | 2,041 (1.8%) |
| **Other/Unknown** | 5,607 (6.3%) | 1,515 (6.4%) | 7,122 (6.3%) |
| **Hospital** |  |  |  |
| **A** | 4,989 (5.6%) | 3,160 (13.3%) | 8,149 (7.2%) |
| **B** | 10,894 (12.3%) | 2,395 (10.1%) | 13,289 (11.8%) |
| **C** | 7,408 (8.3%) | 1,865 (7.8%) | 9,273 (8.2%) |
| **D** | 3,115 (3.5%) | 1,431 (6.0%) | 4,546 (4.0%) |
| **E** | 998 (1.1%) | 740 (3.1%) | 1,738 (1.5%) |
| **F** | 13,86 (1.6%) | 643 (2.7%) | 2,029 (1.8%) |
| **G** | 16,55 (1.9%) | 1,167 (4.9%) | 2,822 (2.5%) |
| **H** | 10,00 (1.1%) | 1,345 (5.7%) | 2,345 (2.1%) |
| **I** | 3,189 (3.6%) | 2,959 (12.4%) | 6,148 (5.5%) |
| **J** | 13,315 (15.0%) | 1,855 (7.8%) | 15,170 (13.5%) |
| **K** | 37,399 (42.1%) | 2,855 (12.0%) | 40,254 (35.8%) |
| **L** | 2,234 (2.5%) | 1,376 (5.8%) | 3,610 (3.2%) |
| **M** | 1,229 (1.4%) | 1,988 (8.4%) | 3,217 (2.9%) |
| **Entry time** |  |  |  |
| **07/2021** | 40,073 (45.1%) | 9,351 (39.3%) | 49,424 (43.9%) |
| **08/2021** | 48,738 (54.9%) | 14,428 (60.7%) | 63,166 (56.1%) |
| **Obesity** |  |  |  |
| **Nonobese** | 27,459 (30.9%) | 10,475 (44.1%) | 37,934 (33.7%) |
| **Obese** | 50,789 (57.2%) | 10,385 (43.7%) | 61,174 (54.3%) |
| **Unknown** | 10,563 (11.9%) | 2,919 (12.3%) | 13,482 (12.0%) |
| **PMCA** |  |  |  |
| **0** | 64,877 (73.1%) | 14,509 (61.0%) | 79,386 (70.5%) |
| **1** | 14,988 (16.9%) | 4,653 (19.6%) | 19,641 (17.4%) |
| **2** | 8,946 (10.1%) | 4,617 (19.4%) | 13,563 (12.0%) |
| **Negative tests prior entry** |  |  |  |
| **0** | 73,356 (82.6%) | 13,309 (56.0%) | 86,665 (77.0%) |
| **1** | 11,450 (12.9%) | 6,220 (26.2%) | 17,670 (15.7%) |
| **2** | 2,611 (2.9%) | 2,213 (9.3%) | 4,824 (4.3%) |
| **>=3** | 1,394 (1.6%) | 2,037 (8.6%) | 3,431 (3.0%) |

**Table 2.** Baseline characteristics of children 5 to 11 and adolescents 12 to 20 years of age in the study of overall effectiveness, direct and indirect effect of the BNT162b2 vaccine on long COVID during the period when the Omicron variant was prevalent.

|  | *Omicron study in children* |  |  | *Omicron study in adolescents* |  |  |
| --- | --- | --- | --- | --- | --- | --- |
|  | Vaccinated<br>(N=101,277) | Unvaccinated<br>(N=87,617) | Overall<br>(N=188,894) | Vaccinated<br>(N=37,724) | Unvaccinated<br>(N=47,011) | Overall<br>(N=84,735) |
| **Age** |  |  |  |  |  |  |
| Median [Q1, Q3] | 8 [6, 10] | 7 [6, 9] | 8 [6, 10] | 15 [13, 17] | 15 [13, 17] | 15 [13, 17] |
| **Distribution** |  |  |  |  |  |  |
| 5 | 14,831 (14.6%) | 18,260 (20.8%) | 33,091 (17.5%) |  |  |  |
| 6 | 14,424 (14.2%) | 15,771 (18.0%) | 30,195 (16.0%) |  |  |  |
| 7 | 14,142 (14.0%) | 13,143 (15.0%) | 27,285 (14.4%) |  |  |  |
| 8 | 14,027 (13.9%) | 11,545 (13.2%) | 25,572 (13.5%) |  |  |  |
| 9 | 14,245 (14.1%) | 10,445 (11.9%) | 24,690 (13.1%) |  |  |  |
| 10 | 14,484 (14.3%) | 9,587 (10.9%) | 24,071 (12.7%) |  |  |  |
| 11 | 15,124 (14.9%) | 8,866 (10.1%) | 23,990 (12.7%) |  |  |  |
| 12 |  |  |  | 8,208 (21.8%) | 6,972 (14.8%) | 15,180 (17.9%) |
| 13 |  |  |  | 5,260 (13.9%) | 6,538 (13.9%) | 11,798 (13.9%) |
| 14 |  |  |  | 4,948 (13.1%) | 6,288 (13.4%) | 11,236 (13.3%) |
| 15 |  |  |  | 4,650 (12.3%) | 6,331 (13.5%) | 10,981 (13.0%) |
| 16 |  |  |  | 4,350 (11.5%) | 5,953 (12.7%) | 10,303 (12.2%) |
| 17 |  |  |  | 4,224 (11.2%) | 4,963 (10.6%) | 9,187 (10.8%) |
| 18 |  |  |  | 2,631 (7.0%) | 3,941 (8.4%) | 6,572 (7.8%) |
| 19 |  |  |  | 1,894 (5.0%) | 3,510 (7.5%) | 5,404 (6.4%) |
| 20 |  |  |  | 1,559 (4.1%) | 2,515 (5.3%) | 4,074 (4.8%) |
| **Gender** |  |  |  |  |  |  |
| Female | 49,080 (48.5%) | 41,313 (47.2%) | 90,393 (47.9%) | 20,617 (54.7%) | 25,109 (53.4%) | 45,726 (54.0%) |
| Male | 52,197 (51.5%) | 46,304 (52.8%) | 98,501 (52.1%) | 17,107 (45.3%) | 21,902 (46.6%) | 39,009 (46.0%) |
| **Ethnicity** |  |  |  |  |  |  |
| White | 22,763 (22.5%) | 35,040 (40.0%) | 57,803 (30.6%) | 9,532 (25.3%) | 22,538 (47.9%) | 32,070 (37.8%) |
| Black/AA | 19,547 (19.3%) | 17,902 (20.4%) | 37,449 (19.8%) | 9,750 (25.8%) | 9,499 (20.2%) | 19,249 (22.7%) |
| Hispanic | 40,242 (39.7%) | 22,020 (25.1%) | 62,262 (33.0%) | 13,506 (35.8%) | 9,389 (20.0%) | 22,895 (27.0%) |
| Asian/PI | 8,136 (8.0%) | 2,904 (3.3%) | 11,040 (5.8%) | 1,734 (4.6%) | 1,153 (2.5%) | 2,887 (3.4%) |
| Multiple | 2,317 (2.3%) | 2,419 (2.8%) | 4,736 (2.5%) | 662 (1.8%) | 920 (2.0%) | 1,582 (1.9%) |
| Other/Unknown | 8,272 (8.2%) | 7,332 (8.4%) | 15,604 (8.3%) | 2,540 (6.7%) | 3,512 (7.5%) | 6,052 (7.1%) |
| **Hospital** |  |  |  |  |  |  |
| A | 5,167 (5.1%) | 8,083 (9.2%) | 13,250 (7.0%) | 2,183 (5.8%) | 4,623 (9.8%) | 6,806 (8.0%) |
| **B** | 14,578 (14.4%) | 10,424 (11.9%) | 25,002 (13.2%) | 5,905 (15.7%) | 5,526 (11.8%) | 11,431 (13.5%) |
| **C** | 6,081 (6.0%) | 5,947 (6.8%) | 12,028 (6.4%) | 1,870 (5.0%) | 3,438 (7.3%) | 5,308 (6.3%) |
| **D** | 3,265 (3.2%) | 4,092 (4.7%) | 7,357 (3.9%) | 1,534 (4.1%) | 3,821 (8.1%) | 5,355 (6.3%) |
| **E** | 582 (0.6%) | 1,502 (1.7%) | 2,084 (1.1%) | 255 (0.7%) | 1,605 (3.4%) | 1,860 (2.2%) |
| **F** | 4,853 (4.8%) | 2,621 (3.0%) | 7,474 (4.0%) | 652 (1.7%) | 1,462 (3.1%) | 2,114 (2.5%) |
| **G** | 1,026 (1.0%) | 2,246 (2.6%) | 3,272 (1.7%) | 662 (1.8%) | 2,409 (5.1%) | 3,071 (3.6%) |
| **H** | 1,200 (1.2%) | 1,727 (2.0%) | 2,927 (1.5%) | 757 (2.0%) | 1,618 (3.4%) | 2,375 (2.8%) |
| **I** | 5,893 (5.8%) | 8,899 (10.2%) | 14,792 (7.8%) | 2,116 (5.6%) | 3,619 (7.7%) | 5,735 (6.8%) |
| **J** | 10,309 (10.2%) | 9,539 (10.9%) | 19,848 (10.5%) | 3,813 (10.1%) | 4,307 (9.2%) | 8,120 (9.6%) |
| **K** | 39,011 (38.5%) | 12,749 (14.6%) | 51,760 (27.4%) | 16,144 (42.8%) | 6,422 (13.7%) | 22,566 (26.6%) |
| **L** | 1,895 (1.9%) | 3,340 (3.8%) | 5,235 (2.8%) | 1,063 (2.8%) | 3,103 (6.6%) | 4,166 (4.9%) |
| **M** | 1,356 (1.3%) | 5,821 (6.6%) | 7,177 (3.8%) | 770 (2.0%) | 5,058 (10.8%) | 5,828 (6.9%) |
| **N** | 433 (0.4%) | 1,142 (1.3%) | 1,575 (0.8%) |  |  |  |
| **O** | 1,626 (1.6%) | 2,555 (2.9%) | 4,181 (2.2%) |  |  |  |
| **P** | 2,700 (2.7%) | 2,936 (3.4%) | 5,636 (3.0%) |  |  |  |
| **Q** | 1,302 (1.3%) | 3,994 (4.6%) | 5,296 (2.8%) |  |  |  |
| **Entry time** |  |  |  |  |  |  |
| **01/2022** | 54,823 (54.1%) | 31,111 (35.5%) | 85,934 (45.5%) | 17,583 (46.6%) | 16,438 (35.0%) | 34,021 (40.1%) |
| **02/2022** | 19,133 (18.9%) | 15,962 (18.2%) | 35,095 (18.6%) | 7,425 (19.7%) | 8,231 (17.5%) | 15,656 (18.5%) |
| **03/2022** | 9,420 (9.3%) | 11,456 (13.1%) | 20,876 (11.1%) | 4,136 (11.0%) | 6,190 (13.2%) | 10,326 (12.2%) |
| **04/2022** | 4,891 (4.8%) | 7,551 (8.6%) | 12,442 (6.6%) | 2,422 (6.4%) | 4,660 (9.9%) | 7,082 (8.4%) |
| **05/2022** | 4,160 (4.1%) | 7,982 (9.1%) | 12,142 (6.4%) | 2,148 (5.7%) | 4,384 (9.3%) | 6,532 (7.7%) |
| **06/2022** | 4,565 (4.5%) | 7,291 (8.3%) | 11,856 (6.3%) | 2,162 (5.7%) | 3,752 (8.0%) | 5,914 (7.0%) |
| **07/2022** | 4,285 (4.2%) | 6,264 (7.1%) | 10,549 (5.6%) | 1,848 (4.9%) | 3,356 (7.1%) | 5,204 (6.1%) |
| **Obesity** |  |  |  |  |  |  |
| **Nonobese** | 34,372 (33.9%) | 38,168 (43.6%) | 72,540 (38.4%) | 11,249 (29.8%) | 19,806 (42.1%) | 31,055 (36.6%) |
| **Obese** | 55,691 (55.0%) | 40,622 (46.4%) | 96,313 (51.0%) | 21,757 (57.7%) | 21,651 (46.1%) | 43,408 (51.2%) |
| **Unknown** | 11,214 (11.1%) | 8,827 (10.1%) | 20,041 (10.6%) | 4,718 (12.5%) | 5,554 (11.8%) | 10,272 (12.1%) |
| **PMCA** |  |  |  |  |  |  |
| **0** | 78,261 (77.3%) | 59,388 (67.8%) | 137,649 (72.9%) | 28,138 (74.6%) | 30,080 (64.0%) | 58,218 (68.7%) |
| **1** | 14,888 (14.7%) | 15,513 (17.7%) | 30,401 (16.1%) | 6,016 (15.9%) | 8,789 (18.7%) | 14,805 (17.5%) |
| **2** | 8,128 (8.0%) | 12,716 (14.5%) | 20,844 (11.0%) | 3,570 (9.5%) | 8,142 (17.3%) | 11,712 (13.8%) |
| **Negative tests prior entry** |  |  |  |  |  |  |
| **0** | 72,618 (71.7%) | 45,099 (51.5%) | 117,717 (62.3%) | 29,406 (78.0%) | 25,020 (53.2%) | 54,426 (64.2%) |
| **1** | 19,079 (18.8%) | 24,973 (28.5%) | 44,052 (23.3%) | 5,833 (15.5%) | 13,486 (28.7%) | 19,319 (22.8%) |
| **2** | 5,893 (5.8%) | 9,905 (11.3%) | 15,798 (8.4%) | 1,561 (4.1%) | 4,750 (10.1%) | 6,311 (7.4%) |
| >=3 | 3,687 (3.6%) | 7,640 (8.7%) | 11,327 (6.0%) | 924 (2.4%) | 3,755 (8.0%) | 4,679 (5.5%) |

### Overall vaccine effectiveness on long COVID

During the Delta variant phase, the BNT162b2 vaccine exhibited an estimated overall vaccine effectiveness against long COVID of 95.4% (95% CI: 90.9% to 97.7%) among adolescents. During the Omicron phase, the estimated overall vaccine effectiveness against long COVID was 60.2% (95% CI: 40.3% to 73.5%) for children. For adolescents, it was 75.1% (95% CI: 50.4% to 87.5%). The BNT162b2 vaccines have shown higher effectiveness during the Delta variant period and are of greater magnitude in adolescents compared to children.

### Direct and indirect effects on long COVID

The direct effect quantifies the vaccine’s impact on long COVID outcomes, separate from its protection against SARS-CoV-2 infection. Conversely, the indirect effect measures the vaccine’s benefit derived from its protective role against the SARS-CoV-2 infection. We refer to Figure S1 of the Supplementary Appendix for a visual representation of the hypothesized effect pathway.

During the phase dominated by the Delta variant, the BNT162b2 vaccine showed an estimated direct effect against long COVID with a relative risk of 1.08 (95% CI: 0.75 to 1.55), which suggests that, outside of its preventive function against SARS-CoV-2 infection, vaccination prior to infection does not significantly modify the risk of long COVID. The corresponding indirect effect was estimated as 0.04 (95% CI: 0.03 to 0.05), underscoring that the benefit of the vaccine in preventing long COVID outcomes is largely attributable to its capacity to mitigate the risk of SARS-CoV-2 infection.

During the Omicron variant phase, for children, the direct effect against long COVID was estimated as 1.24 (95% CI: 0.92 to 1.66), while the indirect effect was estimated as 0.31 (95% CI: 0.23 to 0.42). For adolescents in the same phase, the estimated direct effect against long COVID was 0.91 (95% CI: 0.69 to 1.19), while the indirect effect was 0.21 (95% CI: 0.16 to 0.27). These findings during the Omicron phase align with those from the Delta phase, suggesting that the primary effectiveness of the vaccine in mitigating long COVID is through its protection against the initial risk of SARS-CoV-2 infection.

### Body-system-focused PASC condition clusters

Table 5 presents the estimated overall vaccine effectiveness, along with direct and indirect effects on PASC condition clusters focused on specific body systems, namely cardiac, gastrointestinal, musculoskeletal, respiratory, and syndromic clusters. The findings generally align with the vaccine’s overall effectiveness and mediating effects on the conclusive or probable diagnosis of long COVID.

**Table 3.** Patient follow-up and clinical measures in studying BNT162b2 vaccine on long COVID risks in children and adolescents.

|  | Vaccinated | Unvaccinated | Overall |
| --- | --- | --- | --- |
| *Delta study in adolescents* |  |  |  |
| **Follow-up** |  |  |  |
| Total follow-up — no. of person-wk | 1,521,152 | 397,762 | 1,918,914 |
| Median [Q1, Q3] | 17 [15, 19] | 17 [15, 19] | 17 [15, 19] |
| **Incidence rate per 10,000 person-wk** |  |  |  |
| Documented infection | 1.97 | 53.10 | 12.56 |
| Conclusive or probable PASC | 0.11 | 3.54 | 0.82 |
| Respiratory cluster | 0.01 | 0.63 | 0.14 |
| Musculoskeletal cluster | 0.03 | 0.68 | 0.17 |
| Cardiac cluster | 0.01 | 0.70 | 0.16 |
| Syndromic cluster | 0.03 | 1.26 | 0.29 |
| Gastrointestinal cluster | 0.02 | 0.38 | 0.09 |
| *Omicron study in children* |  |  |  |
| **Follow-up** |  |  |  |
| Total follow-up — no. of person-wk | 4,120,783 | 3,265,395 | 7,386,178 |
| Median [Q1, Q3] | 44 [39, 46] | 40 [31, 45] | 42 [35, 46] |
| **Incidence rate per 10,000 person-wk** |  |  |  |
| Documented infection | 4.56 | 16.43 | 9.81 |
| Conclusive or probable PASC | 0.33 | 1.07 | 0.66 |
| Respiratory cluster | 0.15 | 0.41 | 0.27 |
| Musculoskeletal cluster | 0.04 | 0.15 | 0.09 |
| Cardiac cluster | 0.05 | 0.14 | 0.09 |
| Syndromic cluster | 0.08 | 0.23 | 0.15 |
| Gastrointestinal cluster | 0.06 | 0.15 | 0.10 |
| *Omicron study in adolescents* |  |  |  |
| **Follow-up** |  |  |  |
| Total follow-up — no. of person-wk | 1,491,914 | 1,747,295 | 3,239,209 |
| Median [Q1, Q3] | 43 [36, 46] | 40 [31, 45] | 41 [33, 45] |
| **Incidence rate per 10,000 person-wk** |  |  |  |
| Documented infection | 3.91 | 21.19 | 13.23 |
| Conclusive or probable PASC | 0.24 | 1.43 | 0.88 |
| Respiratory cluster | 0.03 | 0.29 | 0.17 |
| **Musculoskeletal cluster** | 0.05 | 0.27 | 0.17 |
| **Cardiac cluster** | 0.07 | 0.23 | 0.16 |
| **Syndromic cluster** | 0.08 | 0.43 | 0.27 |
| **Gastrointestinal cluster** | 0.01 | 0.16 | 0.09 |

**Table 4.** Estimated overall effectiveness, direct and indirect effects of BNT162b2 vaccines on conclusive or probable diagnosis of long COVID in children and adolescents.

| Vaccine Effectiveness (in %) and 95 CI | Direct Effect | Indirect Effect |
| --- | --- | --- |
| *Delta study in adolescents* |  |  |
| 95.4% (90.9, 97.7) | 1.08 (0.75, 1.55) | 0.04 (0.03, 0.05) |
| *Omicron study in children* |  |  |
| 60.2% (40.3, 73.5) | 1.24 (0.92, 1.66) | 0.31 (0.23, 0.42) |
| *Omicron study in adolescents* |  |  |
| 75.1% (50.4, 87.5) | 0.91 (0.69, 1.19) | 0.21 (0.16, 0.27) |

Vaccine effectiveness: The estimated marginal/total effect of BNT162b2 vaccines on the diagnosis of long COVID in children or adolescents, including both direct and indirect effects.
Direct effect: The estimated effect of BNT162b2 vaccines on the diagnosis of long COVID beyond its effect on the mediator (i.e., documented infection of SARS-CoV-2).
Indirect effect: The estimated effect of BNT162b2 vaccines on the diagnosis of long COVID through the mediator (i.e., documented infection of SARS-CoV-2).

**Table 5.** Estimated overall effectiveness, direct and indirect effects of the BNT162b2 vaccine on body-system-focused PASC condition clusters in children and adolescents.

|  | Vaccine Effectiveness<br>(in %) and 95 CI | Direct Effect | Indirect Effect |
| --- | --- | --- | --- |
| *Delta study in adolescents* |  |  |  |
| *Respiratory cluster* | 97.7% (95.4, 98.9) | 0.78 (0.44, 1.40) | 0.03 (0.01, 0.05) |
| *Musculoskeletal cluster* | 92.9% (86.0, 96.4) | 1.63 (0.88, 3.04) | 0.04 (0.02, 0.07) |
| *Cardiac cluster* | 96.6% (93.3, 98.3) | 0.77 (0.21, 2.74) | 0.04 (0.01, 0.14) |
| *Syndromic cluster* | 96.2% (92.5, 98.1) | 1.16 (0.58, 2.33) | 0.03 (0.01, 0.06) |
| *Gastrointestinal cluster* | 90.2% (80.5, 95.1) | 2.41 (1.47, 3.94) | 0.04 (0.02, 0.06) |
| *Omicron study in children* |  |  |  |
| *Respiratory cluster* | 44.3% (16.5, 62.9) | 1.87 (1.43, 2.45) | 0.29 (0.22, 0.38) |
| *Musculoskeletal cluster* | 71.3% (56.5, 81.1) | 0.49 (0.23, 1.05) | 0.57 (0.27, 1.21) |
| *Cardiac cluster* | 58.3% (36.6, 72.5) | 1.17 (0.41, 3.39) | 0.34 (0.12, 0.97) |
| *Syndromic cluster* | 61.9% (42.2, 74.8) | 1.13 (0.54, 2.37) | 0.33 (0.16, 0.68) |
| *Gastrointestinal cluster* | 46.7% (18.5, 65.2) | 1.63 (0.86, 3.09) | 0.32 (0.17, 0.59) |
| *Omicron study in adolescents* |  |  |  |
| *Respiratory cluster* | 87.4% (74.9, 93.7) | 0.93 (0.61, 1.41) | 0.10 (0.07, 0.16) |
| *Musculoskeletal cluster* | 76.9% (53.8, 88.4) | 0.58 (0.35, 0.95) | 0.30 (0.19, 0.49) |
| *Cardiac cluster* | 27.2% (0.0, 63.5) | 2.72 (1.89, 3.91) | 0.20 (0.14, 0.29) |
| *Syndromic cluster* | 73.3% (46.7, 86.7) | 0.69 (0.42, 1.13) | 0.29 (0.18, 0.48) |
| *Gastrointestinal cluster \** | - | - | - |

\*: Unreliable estimate due to small number of events.

During the phase dominated by the Delta variant, the BNT162b2 vaccine demonstrated consistent effectiveness across different body-system-focused PASC condition clusters, ranging from 90.2% to 97.7%. In the Omicron phase, among children, the BNT162b2 vaccine demonstrated the highest effectiveness against the musculoskeletal cluster at 71.5% (95% CI: 56.5% to 81.1%), which was slightly lower against the respiratory cluster 44.3% (95% CI: 16.5% to 62.9%) and gastrointestinal cluster 46.7% (95% CI: 18.5% to 65.2%). Meanwhile, in the Omicron phase among adolescents, the BNT162b2 vaccine displayed the highest effectiveness against the respiratory cluster at 87.4% (95% CI: 74.9% to 93.7%), and the lowest effectiveness against the cardiac cluster, with an estimate of 27.2% (95% CI: 0.0% to 63.5%).

In the analysis of body-system-focused PASC condition clusters across three study cohorts, the majority of the estimated direct effects did not attain statistical significance, except the gastrointestinal cluster in the Delta phase among adolescents, respiratory cluster in the Omicron phase among children, and cardiac cluster in the Omicron phase among adolescents. The estimated indirect effects indicate that the majority of the vaccine’s protective effect was achieved through protecting against SARS-CoV-2 infection.

### Sensitivity analysis

Sections S5-9 presents the results of sensitivity analyses that were conducted to evaluate the robustness of research findings regarding the study design. Section S5 of the Supplementary Appendix outlines the estimated effects observed during the Delta and Omicron phases when focusing on the adolescent age group of 12 to 17 years. In the sensitivity analysis, which utilized a narrowed observation period within the Delta phase to ensure that the observed long COVID outcomes resulted from infections occurring in that phase, the results presented in Section S6 are consistent with the primary research findings. Moreover, Tables S26, S28, and S30 showcase the percentage of participants who completed a primary series of BNT162b2 vaccines across three different studies. Section S9 summarizes the overall vaccine effectiveness and mediating effects of two-dose vaccination, which yields consistent results with the primary analysis.

Sections S4 and S10-11 the Supplementary Appendix present the results of sensitivity analyses evaluating the impact of statistical methods. Section S10 presents findings on the overall vaccine effectiveness and mediation effects, leveraging a regression-based approach for causal mediation analysis. The results yield reasonably consistent findings as in the primary analysis. In addition, Section S11 details the proximal analysis from three studies, incorporating two negative control exposures and two negative outcomes. After adjusting for the potential of unmeasured confounders through proximal analysis, the results yield statistically insignificant direct effect estimates confirming the findings from the primary analysis that the predominant benefit of the vaccine in protecting against long COVID outcomes arises from its protective effect against SARS-CoV-2 infections.

## Discussion

We identified three study cohorts to assess the effectiveness of the BNT162b2 vaccine on long COVID outcomes using data from a national network of pediatric health systems in the U.S. Utilizing causal mediation analysis, we estimated the vaccine’s overall effectiveness and direct and indirect impacts via specific mediating pathways. Our findings indicated a high overall protective effect of BNT162b2 against long COVID during the period dominated by the Delta variant and moderate effects during the Omicron period. The estimated direct effects suggested that beyond the protective role of vaccination against SARS-CoV-2 infection, pre-infection vaccination does not significantly modify the likelihood of long COVID (i.e., conclusive or probable long COVID as well as body-system-focused PASC condition clusters). The estimated direct and indirect effects indicated that the vaccine’s primary advantage in protecting against long COVID outcomes stems from its ability to reduce the risk of SARS-CoV-2 infection. The higher effectiveness of the BNT162b2 vaccine during the Delta phase, relative to the Omicron period, can be attributed both to its protection against Delta infections and to the fact that the primary benefit of the vaccine on long COVID stems from its capacity to prevent infections.

Our study employed the causal mediation analysis to comprehensively investigate the effectiveness of the BNT162b2 vaccine on long COVID outcomes, which possesses several attractive features compared to existing studies. First, most of the existing studies focused on the vaccine’s effectiveness against long COVID risks within the infected population, while by employing the causal mediation analysis, the overall vaccine effectiveness reported in our study reflected the protective benefits of vaccination to the pediatric population that were both infected and uninfected, holding substantial relevance to public health initiatives. We note that there are a few parallel studies assessing the effectiveness of vaccination against long COVID in the pediatric population^54,55^. Second, in the existing studies, the approach by conditioning on post-treatment variables (i.e., infection) may suffer from the collider bias highlighted in Hernan et al. (2023)^36^, which could produce spurious negative associations. Such an approach could underestimate the true impact of vaccination on long COVID, especially under conditions in which vaccination itself decreases the risk of infection, and thereby, indirectly reduces the risk of developing long COVID. Third, by employing causal mediation analysis to disentangle the vaccine’s impact on long COVID outcomes, the study provides a more nuanced understanding of the role of vaccination and informs decision-making. Our findings on direct effects suggest that documented infections after vaccination could pose a comparable risk of long COVID compared to documented infections occurring without prior vaccination. This highlights the necessity of persistently focusing on preventing SARS-CoV-2 infections as a focus of public health policy to mitigate the risk of long COVID.

Furthermore, our study has several additional strengths. First, to the best of our knowledge, this is the first study of the impact of COVID-19 vaccines on long COVID outcomes that offers insights into both the overall vaccine effectiveness and effects through distinct mediating pathways. Second, considering the complexity of long COVID, the effectiveness against different definitions of long COVID was evaluated: conclusive or probable diagnosis of long COVID through computable phenotype algorithms and PASC condition clusters relevant to body systems. Third, the study cohorts were sourced from a national network of academic medical centers covering a diverse population with broader pediatric demographic, which not only provided a robust sample size but also empowered the detection of rare long COVID outcomes. Fourth, the study included a diverse representation of U.S. pediatric populations from primary care, specialty care, emergency departments, testing centers, and inpatient settings. Fifth, the comprehensive nature of the EHR data enabled us to investigate the effectiveness against diagnosis of long COVID as well as body-system-focused condition clusters while adjusting for a wide array of confounders. Finally, by conducting proximal analysis using negative control variables, our findings offer insights into the impacts of unmeasured confounding variables.

Our study also has several potential limitations. First, effectiveness was investigated in a cohort without previous infection, but potential bias resulting from undocumented infections cannot be ruled out, especially if these occurred differentially in the vaccinated and unvaccinated cohorts. Our inclusion of previous negative COVID-19 tests as a confounder can adjust for the propensity to get tested which could partially adjust for this factor. Second, in the Omicron study involving adolescents, the cohort included adolescents who had their first vaccine after January 1, 2022. Since the use of BNT162b2 vaccines was authorized in adolescents aged 12-15 years on May 10, 2021, this cohort represents a population with late vaccines that may reduce the generalizability of the findings.

Third, vaccine records may be incomplete for individuals who received vaccine doses outside the network’s care delivery sites. To address this, for each study cohort, we only incorporated sites with adequate vaccine data capture, by cross-referencing the EHR data from participating institutions with vaccination statistics from the Centers for Disease Control and Prevention (CDC), which enhances the reliability of the vaccination data in our study. We defined a reference vaccination rate by applying weights to the CDC’s county-level statistics based on the residential addresses of patients. A site was considered to have adequate vaccine data capture if its reported vaccination rate was at least 60% of the CDC’s reference rate.

Fourth, the identification of long COVID in children using EHR data is challenging which may introduce potential bias from inaccurate capturing of outcomes. In this study, besides the ICD10-CM U09.9 diagnosis code of long COVID, we defined probable long COVID using a computable phenotype algorithm, which includes features known to be associated with long COVID in previous statistical studies and chart review validation. However, the possibility of incomplete infection records may decrease the sensitivity of detecting probable long COVID cases using the computable phenotype algorithm. In a prior study, the virus testing data from EHRs were compared against institutional registry data, and the EHRs were found to be more accurate through chart reviews. Additionally, home test-positive patients typically inform hospitals when seeking further medical care. Last, given the rapid evolution of the pandemic, an updated analysis is desirable in the future to assess the findings in the context of the current circulating variants.

In summary, this study, based on national pediatric cohorts in the U.S., revealed protective effectiveness of the BNT162b2 vaccine on long COVID as well as body-system-focused PASC condition clusters. The findings suggested the vaccine’s predominant benefit in protecting against long COVID outcomes is its capacity to reduce the risk of SARS-CoV-2 infection. This study profoundly enriches our understanding of the BNT162b2 vaccine’s impact on long COVID risks within the U.S. pediatric demographic, emphasizing the critical role of vaccination and infection prevention in public health policymaking.

## Disclaimer

This content is solely the responsibility of the authors and does not necessarily represent the official views of the RECOVER Initiative, the NIH, or other funders.

## Data Availability

All data produced in the present study are available upon reasonable request to the authors

## Acknowledgements

Reference NIH Involvement

This study is part of the NIH Researching COVID to Enhance Recovery (RECOVER) Initiative, which seeks to understand, treat, and prevent the post-acute sequelae of SARS-CoV-2 infection (PASC). For more information on RECOVER, visit https://recovercovid.org/

Representative Acknowledgement:

We would like to thank the National Community Engagement Group (NCEG), all patients, caregivers, and community representatives, and all the participants enrolled in the RECOVER Initiative. We also would like to thank Drs. Margot I. Gage Witvliet and Megan L. Fitzgerald for their helpful discussions.

